# Wide application of minimally processed saliva on multiple RT-qPCR kits for SARS-CoV-2 detection in Indonesia

**DOI:** 10.1101/2021.04.01.21254743

**Authors:** Caroline Mahendra, Maria Mardalena Martini Kaisar, Suraj Rajan Vasandani, Sem Samuel Surja, Enty Tjoa, Febie Chriestya, Kathleen Irena Junusmin, Tria Asri Widowati, Astrid Irwanto, Soegianto Ali

**Author notes:** Correspondence: Soegianto Ali, School of Medicine and Health Sciences, Atma Jaya Catholic University of Indonesia, Pluit Raya No. 2, Jakarta 14440, Indonesia. These authors have contributed equally to this work and share first authorship (Caroline Mahendra, Maria Mardalena Martini Kaisar, Suraj Rajan Vasandani).

## Abstract

Saliva as a sample matrix has been an attractive alternative for the detection of SARS-CoV-2. However, due to potential variability in collection and processing steps, evaluating a proposed workflow amongst the local population is recommended. Here, we aim to validate the collection and treatment of human saliva as a direct specimen for RT-qPCR-based detection of SARS-CoV-2 in Indonesia. We demonstrated that SARS-CoV-2 target genes were detected in saliva specimens and remained stable for five days refrigerated or room temperature storage. The method of processing saliva specimens described in this report bypasses the need for an RNA-extraction process, thereby reducing the cost, time, and manpower required for processing samples. The developed method was tested across nine commercial kits, including the benchmark, to demonstrate its wide applicability on multiple existing workflows. Our developed method achieved 86% overall agreement rate compared to paired nasopharyngeal and oropharyngeal swab specimens (NPOP). With the assistance of a saliva sampling device, the collection was found to be more convenient for individuals and improved the overall agreement rate to 97%.

## 1. Introduction

COVID-19 case was first reported to the World Health Organization (WHO) on December 31, 2019, and was declared a pandemic on March 11, 2020 (Timeline of WHO’s response to COVID-19). COVID-19 is caused by SARS-CoV-2 viral infection, an enveloped virus with single-stranded positive-strand genomic RNA (Brian and Baric, 2005). One of the modalities used to diagnose COVID-19 is nucleic acid amplification test (NAAT), including Reverse Transcription quantitative Polymerase Chain Reaction (RT-qPCR), which commonly targets the envelope (E), nucleocapsid (N), RNA-dependent RNA polymerase (RdRP), and spike (S) genes (World Health Organization, 2020). Specimens that could be used for NAAT include those obtained from the upper and lower respiratory tracts and gastrointestinal tracts (Wang et al., 2020). Specimens commonly collected from the upper respiratory tracts are nasopharyngeal and oropharyngeal (NPOP) swabs, and those obtained from the lower respiratory tract include bronchoalveolar lavage. The rate of detecting positive infection from bronchoalveolar lavage is superior to that of NPOP swabs; however, it is commonly obtained from inpatients with severe illness or those undergoing mechanical ventilation (Wang et al., 2020). The stool has also been used as a specimen for NAAT methods for SARS-CoV-2 detection, including among children (Zhang et al., 2020).

Collecting NPOP specimens require trained healthcare personnel and could induce aerosolization that increases the risk of infection to healthcare personnel. Adequate Personal Protective Equipment (PPE) is needed, and specimen collection can only be done in designated sites to reduce the risk of transmission during the procedure. Indonesia is a very vast country with varying levels of access to collection sites at medical facilities. Patients with suspected infections located far from urban facilities may need to travel on public transport for a period of time to arrive at medical centers for NPOP collection (COVID-19 developments in Indonesia). Such situations could increase the risk of disease spread from the individuals’ exposure to the mass population during his/her travel.

Other studies have shown that saliva can serve as an alternative specimen for NAAT-based SARS-CoV-2 detection (Hung et al., 2020; Wyllie et al., 2020). Its non-invasive nature reduces the level of discomfort experienced when sampling, minimizes production of aerosols and does not require a trained healthcare provider, which could allow for flexibility of sampling at various collection sites, including at-home. Although there is a reduction in sensitivity to detecting SARS-CoV-2 from saliva specimens, its specificity remained at par with NPOP specimens, suggesting that saliva is still a reliable specimen (Griesemer et al., 2020; Landry et al., 2020; Xu et al., 2020).

The standard protocol for detecting SARS-CoV-2 using the RT-qPCR method from NPOP swabs requires various consumables. The NPOP swab should be immersed in Viral Transport Medium (VTM) for transportation from the collection site to the laboratory and maintained in cold condition. In the laboratory, the VTM containing NPOP specimens are extracted to isolate viral RNA. This step generally utilizes a commercial RNA extraction kit, which could take up to 1 – 1.5 hours to complete. Purified viral RNA then will be used as a template for RT-qPCR amplification which takes 2 – 3 hours from reaction set up to complete. The whole procedure could take 3 – 4.5 hours from sample collection to result reporting.

Previous reports have demonstrated comparable results of performing RT-qPCR directly from NPOP without the RNA extraction step (Alcoba-Florez et al., 2020; Smyrlaki et al., 2020). Direct PCR omits the need for RNA extraction kits and reduces the turnover time by 1 – 1.5 hours. Currently, in Indonesia, most RNA extraction kits are imported, and particularly during this pandemic, it can be scarce.

Considering the above-mentioned possibilities, we tested and validated the detection of SARS-CoV-2 by RT-qPCR from minimally processed saliva specimens. The validated method would be more convenient for patients, safer for healthcare providers and reduce the time and cost of the current RT-qPCR test to detect COVID-19 infection.

## 2 Materials and Methods

### 2.1 Ethical Clearance

The collection of clinical specimens, NPOP swabs, and saliva specimens were approved by the Institutional Review Board of the School of Medicine and Health Sciences, Atma Jaya Catholic University of Indonesia (No:16/11/KEP-FKIKUAJ/2020).

### 2.2 Study Recruitment

Recruitment for study participants was done in collaboration with multiple SARS-CoV-2 testing sites. Participants were verbally informed about the study and the procedures involved. Written informed consent was obtained from all participants prior to specimen collection. Inclusion criteria for this study were patients who tested positive up to 14-days before specimen collection or were in close contact with a known positive patient. Exclusion criteria were patients who were critically ill, unconscious, and/or intubated.

### 2.3 Specimen Collection

Collection of NPOP swabs and saliva specimens were performed for every patient and within one hour of each other. NPOP swabs were collected by a trained medical professional by inserting separate swabs into the participants’ nasopharyngeal (NP) and oropharyngeal (OP) cavity and immersed into a single tube containing VTM. Prior to saliva specimen collection, patients were required to satisfy a 30-minute fasting period during which they were prohibited from eating, drinking, smoking, tooth brushing, using mouthwash, and other activities that involved the oral cavity. Approximately 2-5 mL of unstimulated saliva was collected into a 50 mL tube without the addition of buffers or any other stabilizing medium. Specimens were then kept on ice during transport and processed in an enhanced Biosafety Laboratory Level 2 facility at the School of Medicine and Health Sciences, Atma Jaya Catholic University of Indonesia.

### 2.4 Viral RNA Extraction

Viral RNA was extracted with QIAamp Viral RNA Mini kit (Catalog # 52906, Qiagen, Hilden Germany) following instructions provided by the manufacturer. In brief, 140µL of VTM containing NPOP swabs or saliva specimens was mixed with lysis buffer, bound to silica membrane present in the spin column, washed twice, and eluted (60µL) as pure RNA.

### 2.5 Viral Nucleic Acid Detection with RT-qPCR

Da An Gene’s Detection Kit for 2019 Novel Coronavirus (2019-nCoV) RNA (PCR-Fluorescence Probing) (Catalog # DA-930) was used as the reference nucleic acid detection kit in this study. The RT-qPCR master mix was prepared following the manufacturer’s recommended instruction - 17µL of PCR reaction solution A and 3µL of PCR reaction solution B for each reaction. The template (5µL) used was either extracted viral RNA (section 2.4), or RNA-extraction-free treated saliva specimens (section 2.9). Upon template addition, strip tubes were briefly spun down to ensure that all liquid is positioned at the bottom of the tube. Thermocycling conditions were as follows: 15 minutes at 50°C, 15 minutes at 95°C, and 45 cycles of 15 seconds at 94°C and 45 seconds at 55°C. The amplification, detection, and analysis were performed using the CFX96 Touch Real-Time PCR detection system (Bio-Rad laboratories). Negative and positive controls were included in each RT-qPCR run. Cycle threshold (Ct) values were analyzed using CFX Maestro software (Bio-Rad laboratories). The Ct-value results represent the amplification cycle in which the fluorescence signal level exceeds the background fluorescence, reflecting the presence of SARS-CoV-2 RNA in the specimen tested. Specimens were interpreted as positive if the cycle threshold (Ct) values for N-gene and ORF1ab were less than 40, and the curve displayed apparent amplification in the typical “S” shaped form. Specimens with no amplification (N/A) or Ct values > 40 for both genes but had internal control amplified were interpreted as negative. Specimens that only had amplification in one of the target genes, N-gene or ORF1ab but not both, or no amplification at all across all the channels, were interpreted as invalid in this study.

### 2.6 Viability of Saliva as a Sample Matrix

Paired NPOP and saliva specimens were collected and used to show the viability of saliva as a sample matrix to detect SARS-CoV-2 RNA (n=116). Viral RNA from both specimens was extracted with the QIAamp Viral RNA Mini kit (section 2.4). Detection of viral RNA from both specimen types was done on our reference kit (section 2.5).

### 2.7 Optimization of RNA-extraction free treatment for saliva specimens

Optimization of RNA-extraction-free treatment of saliva specimens was performed on six previously diagnosed specimens, consisting of positives (n=3) and negatives (n=3). Viral RNA was extracted from NPOP and saliva specimens (section 2.4) before subjected to RT-qPCR to detect SARS-CoV-2 (section 2.5) (**Figure 1A**, Treatments 1 and 2). The remaining saliva specimens were vortexed, aliquoted as 100µL into six 1.5mL microtubes, and subjected to the different RNA-extraction-free treatments (**Figure 1A**, Treatments 3 to 8) before added as templates into the RT-qPCR reaction for viral RNA detection (**Supplementary 1)**(section 2.5).

**Figure 1.**
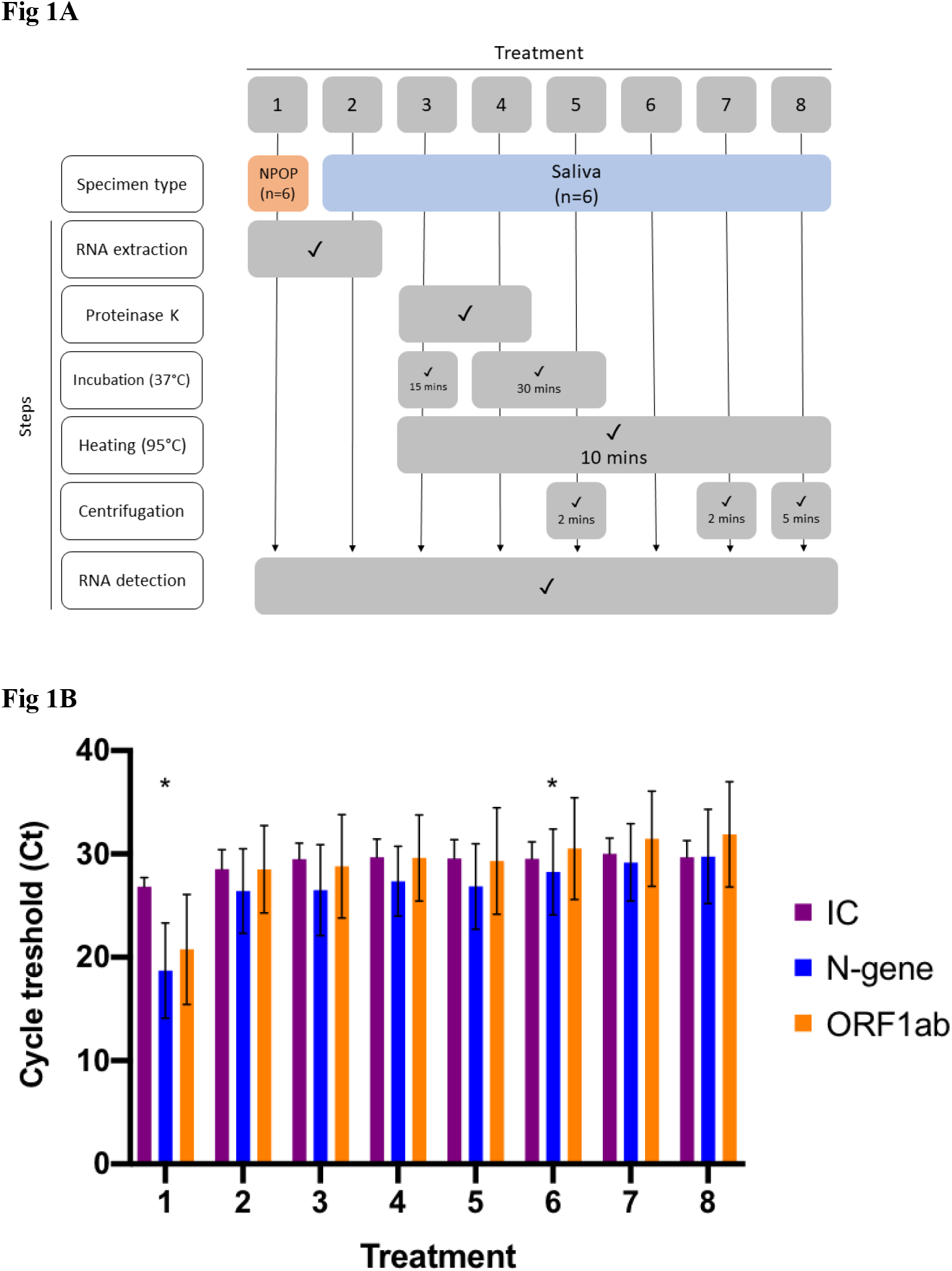
Optimization of RNA-Extraction-free treatment. (A) Flowchart of eight different sample treatments prior SARS-CoV-2 detection (B) Bar graph represents means ± SEM of three SARS-CoV-2 positive specimens tested for each sample treatment. Asterisk (*) denotes the treatments for comparison, where 1 is the comparative method while 6 is the candidate method.

The effectiveness of RNA-extraction-free treatment of saliva specimens was evaluated by comparing Ct values on positive saliva specimens (n=55) that were subjected to RNA extraction Treatment 2 and optimized RNA-extraction-free Treatment 6 (**Figure 1A, Supplementary 1)**

### 2.8 Saliva Specimen Stability for RNA-Extraction-free Treatment

Saliva specimens with Ct values ranging from 14.24 to 32.85 for N-gene and 18.15 to 35.18 for ORF1ab were monitored for specimen stability (n=14). Collected saliva specimens were aliquoted into 100µL in 1.5mL microtubes and stored at room temperature (∼25°C) or inside a refrigerator (2-8°C). Specimens stored at room temperature were tested daily for five days, while those stored in the fridge were tested on days three through five. A tube from each storage condition was retrieved for each time point, and RNA-extraction-free treatment of saliva specimens (section 2.9) was carried out before subjecting specimens to RT-qPCR (section 2.5).

### 2.9 Validating RNA-extraction-free Treatment of Saliva Specimens

Paired NPOP and saliva specimens were collected and used to assess the performance of our RNA-extraction-free treatment of saliva specimens (Treatment 6; **Figure 1A, Supplementary 1**) against extracted viral RNA from NPOP specimens (n=125). Collected specimens arriving in the laboratory were stored in a refrigerator at approximately 2-8°C and processed within 5 days of the collection date. Viral RNA from NPOP swabs and treated saliva were then used as RT-qPCR templates (section 2.5).

### 2.10 Compatibility with Other SARS-CoV-2 RT-qPCR Detection Kits

We assess the compatibility of this method with eight more commercial kits other than the reference, following instructions provided by each manufacturer. Details of genes targeted, internal control, the limit of detection, the number of cycles, cycle threshold cut-off, and template volume are summarized in **Supplementary 2**. Evaluation of each kit was done on specimens previously characterized by our reference kit (n=10 to 13). The samples selected as a pool to subject to different kits should contain at least two positives. Positive samples selected had target genes detected with Ct range from 17.07 to 35.54 for N-gene and 18.48 to 37.14 for ORF1ab.

For kits that required internal control to be added into specimens prior to extraction (Fosun and Maccura), the step was modified to add internal control into the RT-qPCR master mix at 0.1x of its recommended volume. The amplification, detection, and analysis were performed using the CFX96 Touch Real-Time PCR detection system and CFX Maestro software (Bio-Rad laboratories).

### 2.11 Implementation of a Saliva Collection Device

Paired NPOP swab and saliva specimens were collected from collaborating sites during the implementation of the saliva collection device, QuickSpit™ (n=306). NPOP swab specimens were collected and underwent RNA extraction following standard procedures performed by the collaborating sites. NPOP results obtained were used as the benchmark for this experiment. Saliva specimens were collected at 0.5-1.0 mL with QuickSpit™ following instructions for use and processed with RNA-extraction-free treatment before being subjected to RT-qPCR (section 2.5).

### 2.12 Data Management and Statistical Analysis

Collected respondent’s information and RT-qPCR data were stored in a Microsoft Excel database with restricted sharing to authors only. Two-by-two contingency matrices were used to compare PCR results from extracted NPOP and test conditions: extracted saliva (section 2.6), treated saliva (section 2.9), and treated saliva collected with QuickSpit (section 2.12). Only specimens that returned positive or negative results on both conditions were included in calculating agreement rates. Results were reported as overall agreement, positive percent agreement (PPA), and negative percent agreement (NPA), each with 95% scores of confidence interval (95% CI) calculated using the following formulas:

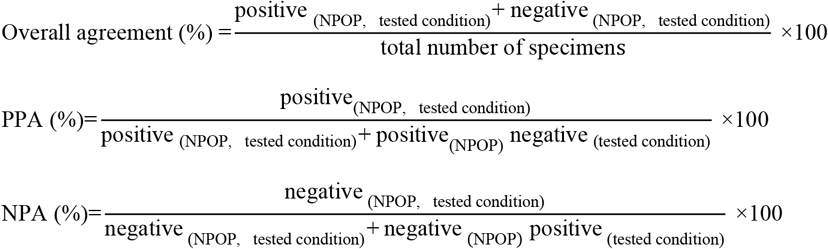

Paired two-tailed t-tests were performed to compare the means of Ct value between different saliva specimen treatments (section 2.7) and between the mean difference of the varying saliva storage conditions (section 2.8). The null hypothesis stated there is no difference in means of Ct value across the tested conditions. Differences with a *p-value* < 0.05 were considered to be statistically significant. Statistical analysis and data visualization were performed using GraphPad Prism version 8 (GraphPad Software, La Jolla, CA USA) and Microsoft Excel for Windows.

The compatibility of the developed method with different RT-qPCR kits was assessed by comparing results obtained on the evaluated kits to those obtained on the reference kit. Results were reported as the rate of invalid specimens, overall agreement, and estimated sensitivity calculated with the following formulas:

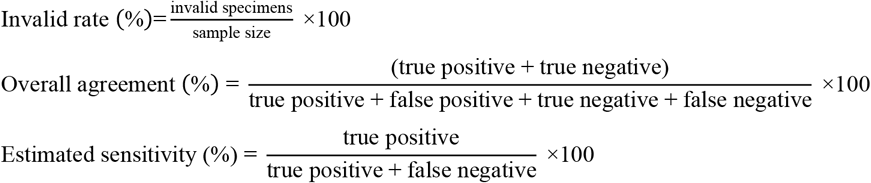

True positives or negatives were defined as specimens that had concordant results on the evaluated and reference kit. False results were defined as specimens that had discordant results – specimens that were positive on the evaluated kit but negative on reference were interpreted as false positives, while specimens that were negative on the evaluated kit but positive on reference were interpreted as false negatives. The specimens that did not return a result on the evaluated kit were defined as invalid.

## 3 Results

### 3.1 Transitioning from NPOP swab to saliva for detection of SARS-CoV-2 by RT-qPCR

We first sought to validate saliva as a specimen suitable for the detection of SARS-CoV-2 by RT-qPCR. A total of 103 samples returned positive or negative results on both specimens out of the collected 116 samples. We found an 89.3% overall agreement between specimens extracted from saliva and NPOP, consisting of 84.5% positive percent agreement (PPA) and 100% negative percent agreement (NPA) (**Table 1, Supplementary 3**). Eleven specimens tested negative on saliva but positive on NPOP. This shows the viability of saliva specimens in detecting SARS-CoV-2.

**Table 1.** SARS-CoV-2 detection from RNA extracted from NPOP *versus* saliva sample types.

| Extracted saliva, n | NPOP |  | Total |
| --- | --- | --- | --- |
|  | Positive | Negative |  |
| Positive | 60 | 0 | 60 |
| Negative | 11 | 32 | 43 |
| Total | 71 | 32 | 103 |
| Agreements | % |  | 95% CI |
| Overall* | 89.32 |  | 81.88-93.93 |
| Positive** | 84.51 |  | 74.35-91.12 |
| Negative*** | 100.00 |  | 89.28-100.00 |

\*Overall agreement = ((60+32)/103) x 100
\*\*Positive agreement = (60/71) x 100
\*\*\*Negative agreement = (32/32) x 100

### 3.2 Development of an RNA-extraction-free treatment of saliva for detection of SARS-CoV-2

To streamline sample treatment for downstream RT-qPCR application, we explored several treatment methods on saliva specimens. This included saliva undergoing heating, the addition of Proteinase K, and concentrating by centrifugation paired with RNA extracts from NPOP and saliva as templates for RT-qPCR reaction (**Figure 1A, Supplementary 1**). We found all treatment methods on saliva specimens resulted in the same qualitative outcome on positive specimens and comparable Ct values for the two target genes (**Figure 1B**).

Given the need for an affordable and scalable yet effective method, we developed a specimen treatment that only involved the heating of saliva (**Figure 1A**, Treatment 6). Paired t-test analysis was done between Ct values obtained from positive specimens extracted from NPOP swabs, Treatment 1, and RNA-extraction-free treatment of saliva specimens, Treatment 6. There was no significant difference in detection of N-gene (*p-value* = 0.102) and ORF1ab (*p-value* = 0.107), demonstrating effectiveness of SARS-CoV-2 detection from heat-treated saliva specimen as compared to RNA extract from NPOP swabs. This method also confirmed qualitative results on negative specimens (**Supplementary 4**).

We generated a linear regression from positive saliva specimens to demonstrate the effectiveness of RNA-extraction-free treatment of saliva specimens, Treatment 6, compared to those subjected to RNA extraction, Treatment 2 (n=55). A strong positive correlation in Ct value for N-gene (coefficient = 1.00, R^2^ = 0.929) and ORF1ab (coefficient = 0.966, R^2^ = 0.837) was observed demonstrating that the Treatment 6 is effective for detection of viral RNA in saliva (**Figure 2**).

**Figure 2.**
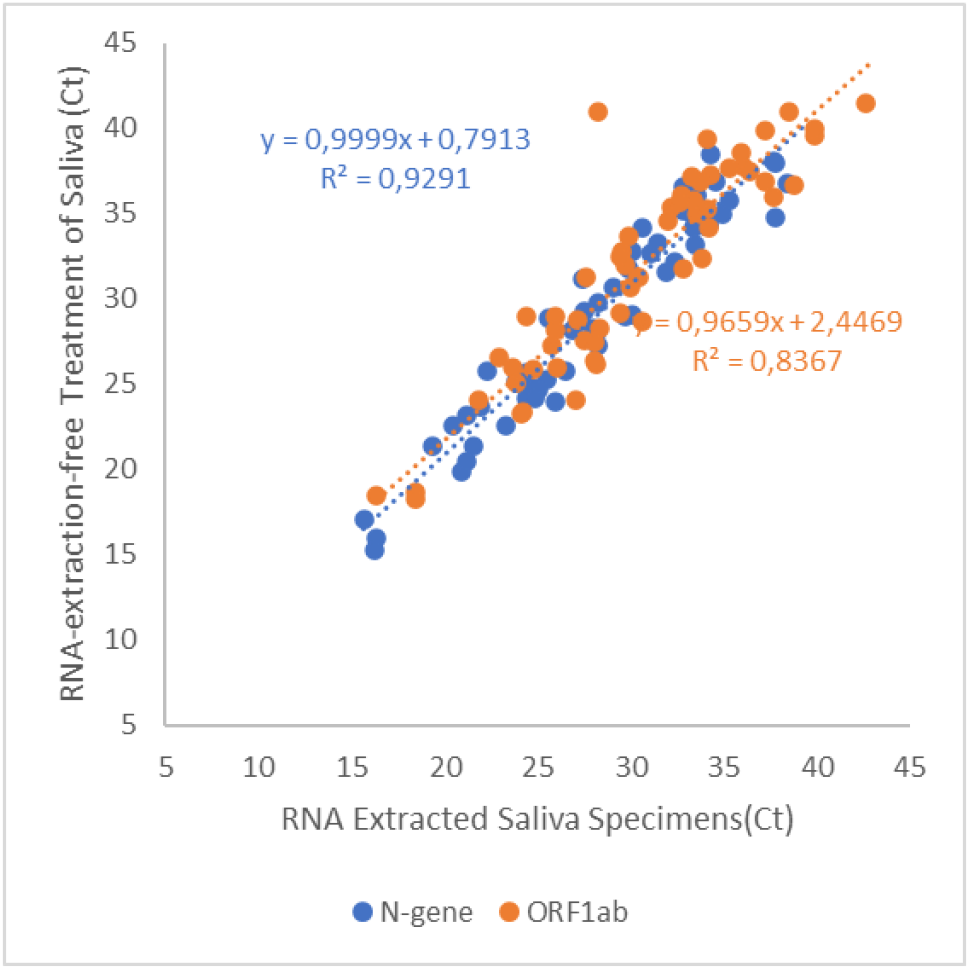
Linear regression of Ct values obtained from adding RNA extracted or heated-only saliva for RT-qPCR template. Data points in blue indicate Ct values for the detection of N-gene while orange for the detection of ORF1ab. Results obtained from extracted RNA from saliva sepcimens were plotted on the x-axis, while heated-only saliva on the y-axis. The line of best fit was plotted for both target genes to display direct proportion between the two variables.

### 3.3 SARS-CoV-2 in saliva specimens remained stable at 4°C and room temperature (RT)

To determine the storage condition for saliva specimens that maintain effective treatment and detection of SARS-CoV-2, we monitored specimen stability at two temperatures on previously confirmed positive saliva specimens for five days (n=14). Each specimen was then subjected to heat treatment followed by RT-qPCR detection at selected time points during the period of storage. We found that detection of both target genes remained stable for 5 days at both storage conditions for specimens from low to high viral load with initial Ct ranging from 14 to 35 (**Figure 3, Supplementary 5**).

**Figure 3.**
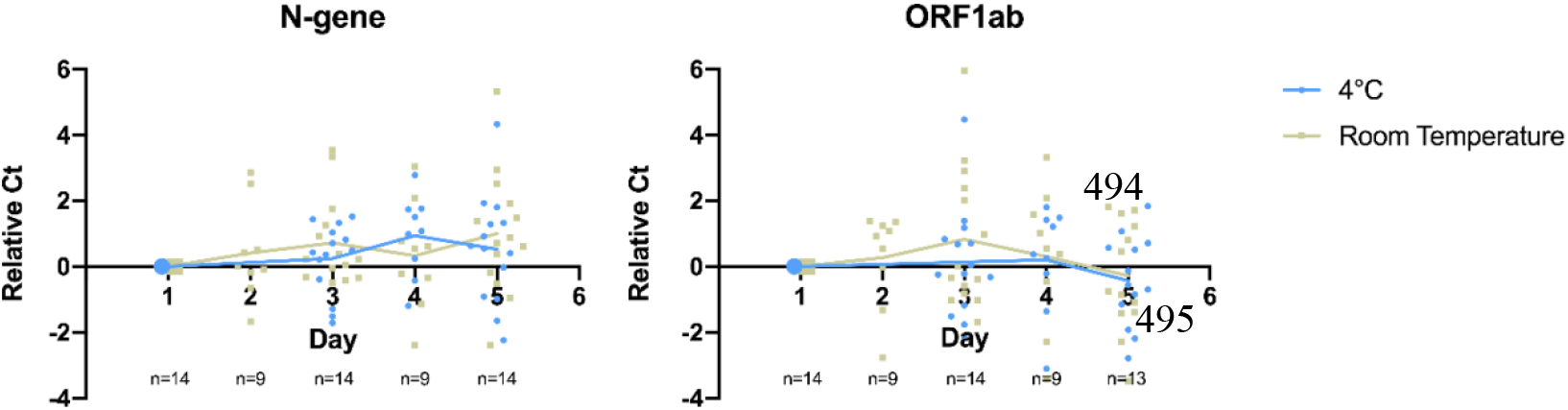
Detection of SARS-CoV-2 target genes N-gene and ORF1ab remained stable for 5 days at cold and room temperature storage conditions. Relative Ct on the y-axis plots the difference between Ct value obtained at a given day and Day 1 for the monitored sample. Data points in blue refer to samples stored inside a refrigerator (2-8°C), while grey refer to samples stored at room temperature. Straight lines display the average relative Ct obtained across storage days.

On detection of N-gene, we found no significant difference in Ct values upon storage for 5 days at 4°C (ΔCt = 0.52, *p-value* = 0.262) and room temperature (ΔCt = 1.00, *p-value* = 0.066). There was no significant difference in the detection of N-gene between storing specimens at both temperatures (*p-value* = 0.341). ORF1ab detection slightly improve upon storing for a period of 5 days, as seen in the decrease in Ct value at 4°C (ΔCt = -0.386, *p-value* = 0.295) and room temperature (ΔCt = -0.281, *p-value* = 0.559), although there was no significant difference between the two temperatures (*p-value =* 0.671). This shows that SARS-CoV-2 remain stable in saliva specimens for up to five days on both storage conditions.

### 3.4 RNA-extraction-free treatment of saliva specimens is reliable for detecting SARS-CoV-2

To assess the performance of the developed method, patients were recruited with written informed consent for collection of NPOP swabs and saliva at the same time point by health care workers. A total of 110 samples returned positive or negative results on both specimens out of the collected 125 samples. NPOP specimens were subjected to QIAamp Viral RNA Extraction, while saliva specimens were treated under the RNA-Extraction-free method, and both were applied as a template for subsequent RT-qPCR detection. We found the overall percent agreement between RNA extract from NPOP swabs and treated saliva specimens to be 86.4%, with 79.2% PPA and 100% NPA (**Table 2, Supplementary 6**). Fifteen specimens tested different results across the two specimen types. This was expected due to variation in specimen types but did not rule out the validity of this method of saliva treatment.

**Table 2.**
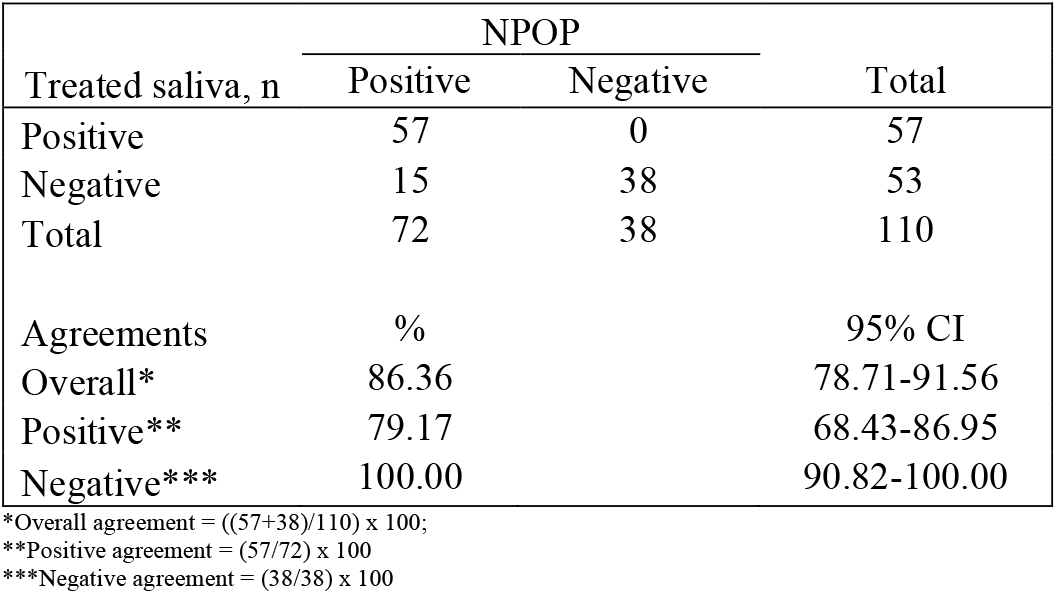
Validation of heated-only saliva as RNA-extraction free treatment.

| Treated saliva, n | NPOP |  | Total |
| --- | --- | --- | --- |
|  | Positive | Negative |  |
| Positive | 57 | 0 | 57 |
| Negative | 15 | 38 | 53 |
| Total | 72 | 38 | 110 |
| Agreements | % |  | 95% CI |
| Overall* | 86.36 |  | 78.71-91.56 |
| Positive** | 79.17 |  | 68.43-86.95 |
| Negative*** | 100.00 |  | 90.82-100.00 |

\*Overall agreement = ((57+38)/110) x 100;
\*\*Positive agreement = (57/72) x 100
\*\*\*Negative agreement = (38/38) x 100

### 3.5 Heat-treated saliva specimen is versatile with a number of SARS-CoV-2 RT-qPCR kits

To assess the versatility of the developed method for saliva treatment, we subjected the heat-treated saliva specimens (n= 10 to 13) for detection using eight more commercial SARS-CoV-2 RT-qPCR kits (**Supplementary 2**). Results obtained were then compared to the reference kit that this treatment method was developed on, Da An Gene. The parameters of interest to assess compatibility of the treatment with a commercial kit were their performance at generating invalid results and overall agreement to results produced by the reference kit.

Most of the kits tested generated invalid rates of under 10%, demonstrating that the treatment did not risk inhibiting reactions of other commercial kits, except for Vazyme which had 40% invalid rates (n=4/10). Two kits obtained results that were in 100% agreement to the reference, Maccura (n=11/11) and Fosun (n=11/11). Ardent (n=10/11) and SD Biosensor (n=11/12) displayed above 90% agreement, followed by Biosewoom at 80% (n=8/10) and Fortitude 70% (n=7/10). 3S and Vazyme had the lowest agreement to the reference, 40% (n=4/10) and 33% (n=2/6) respectively (**Table 3**). Upon closer inspection, the lower overall agreement generated by 3S and Vazyme (n=4/6) were contributed from detection of false positives (3S n=6/10; Vazyme n=4/6; **Supplementary 7**). This could arise due to their limit of detection being lower than the reference kit, resulting in samples of lower viral load detectable by 3S and Vazyme but not by Da An Gene (**Supplementary 2**). Together, our results demonstrated that most kits were compatible with the developed method for treatment of saliva with varying performance, indicating that validation is recommended before implementing the method with any existing workflow.

**Table 3.**
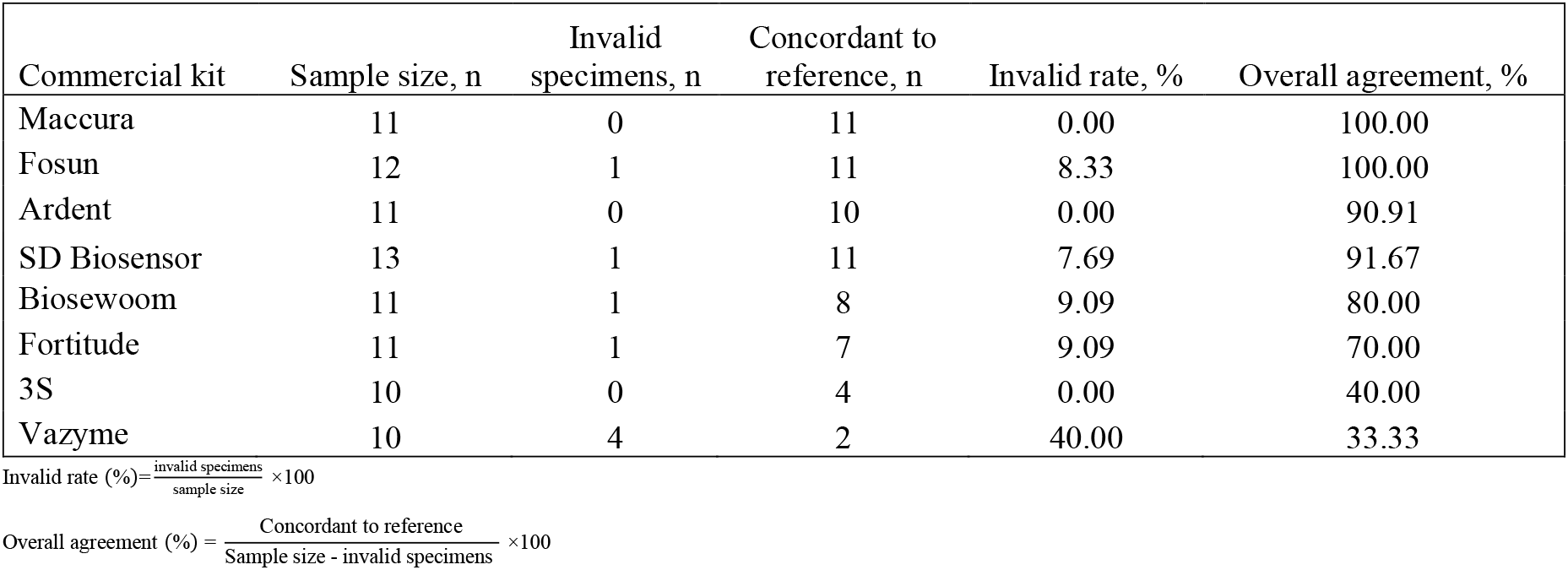
Performance of commercial kits using treated saliva as a template for RT-qPCR. Rates of invalid and overall agreement were calculated as compared to results from the reference, Detection Kit for 2019 Novel Coronavirus (2019-nCoV) by Da An Gene Co.

| Commercial kit | Sample size, n | Invalid specimens, n | Concordant to reference, n | Invalid rate, % | Overall agreement, % |
| --- | --- | --- | --- | --- | --- |
| Maccura | 11 | 0 | 11 | 0.00 | 100.00 |
| Fosun | 12 | 1 | 11 | 8.33 | 100.00 |
| Ardent | 11 | 0 | 10 | 0.00 | 90.91 |
| SD Biosensor | 13 | 1 | 11 | 7.69 | 91.67 |
| Biosewoom | 11 | 1 | 8 | 9.09 | 80.00 |
| Fortitude | 11 | 1 | 7 | 9.09 | 70.00 |
| 3S | 10 | 0 | 4 | 0.00 | 40.00 |
| Vazyme | 10 | 4 | 2 | 40.00 | 33.33 |

### 3.6 Implementation of saliva collection and treatment method for SARS-CoV-2 diagnostics

To better assist with the saliva specimen collection process, we utilized the QuickSpit™ Collection Kit for an easy and safe sampling of human saliva. The study recruitment was continued at multiple collaborating sites using the QuickSpit™ Collection kit. Paired NPOP swabs and saliva were collected and processed to evaluate the percent agreements between extracted RNA from NPOP specimens and RNA-extraction-free treatment of saliva specimens collected with QuickSpit™. A total of 293 samples returned positive or negative results on both specimens out of the collected 306 samples (**Table 4, Supplementary 8**). The use of a collection device improved the agreement between treated-saliva and extracted-NPOP methods of SARS-CoV-2 treatment before RT-qPCR detection, where 90% overall agreement was observed with PPA at 85% and NPA at 100%.

**Table 4.** Implementation of QuickSpit™ as collection and treatment method of saliva specimen.

| Treated saliva collected with QuickSpit™, n | NPOP |  | Total |
| --- | --- | --- | --- |
|  | Positive | Negative |  |
| Positive | 45 | 0 | 45 |
| Negative | 8 | 240 | 248 |
| Total | 53 | 240 | 293 |
| Agreements | % |  | 95% CI |
| Overall* | 97.27 |  | 94.71-98.61 |
| Positive** | 84.91 |  | 72.95-92.15 |
| Negative*** | 100.00 |  | 98.43-100.00 |

\*Overall agreement = ((45+240)/293) x 100;
\*\*Positive agreement = (45/53) x 100
\*\*\*Negative agreement = (240/240) x 100

## 4 Discussion

Numerous references have reported the use of human saliva as an attractive specimen for detection of SARS-CoV-2 infection for its practicality in sampling and processing (Ott et al.; Azzi et al., 2020; Iwasaki et al., 2020; Vogels et al., 2020; Watkins et al., 2020; Wyllie et al., 2020; Xu et al., 2020). Utilizing saliva specimens allows for convenient self-collection without the need for a medical professional inserting swabs into a patient’s nasopharyngeal and/or oropharyngeal cavity. In this study, we validated the application of human saliva as a candidate specimen for detecting SARS-CoV-2, achieving 90% agreement with the current conventional specimens - NPOP swabs. Although some discordance was observed between NPOP swabs and saliva specimens, this is in conjunction with previous reports that some variation exists between specimen types (Griesemer et al., 2020; Landry et al., 2020; Vogels et al., 2020; Xu et al., 2020). This could arise from individuals bearing the virus only in their nasopharyngeal cavity but not in their oral cavity. NPOP swab collects specimen from both respiratory tract sites into a single VTM-containing tube, whereas saliva specimens only collect from the oral cavity. Further investigation on the viral pathway of infection is needed to understand these cases of discordance between specimen types. Results obtained in this study still confirm that saliva is a viable alternative specimen for SARS-CoV-2 detection.

Our developed method could increase the scale of SARS-CoV-2 testing since the treatment requires less manpower and time. Furthermore, minimally processed saliva specimens reduce the cost of testing by two-folds: 1) exempting the cost of trained professionals for specimen collection and the need of VTM and swabs, and 2) removing the need for the RNA extraction process. Saliva specimen collection could be performed at home and sent to the laboratory in a safe box, hence omitting the need for the suspected person to travel to the diagnostic center and, thereby, reduce the chances of transmission that could occur during the journey.

The invalid rate for testing saliva was 2% higher than NPOP specimens (invalid specimens in saliva = 7.8% (n=116); NPOP = 5.1% (n=564)) (**Supplementary 9**). This could arise from the carryover of interfering substances from the oral cavity during collection, which was observed in some specimens that were colored, viscous, and/or particulate. Such substances are also potential causes of reaction inhibitors, as seen in the increase of invalid rates from 7% (n=116) in extracted saliva to 8.8% (n=125) with treated saliva. The use of a saliva collection device reduced invalid rates on saliva specimens to 3.6% (n=306) and improved agreements between treated-saliva and extracted NPOP for viral detection. Overall improvement was seen at an increase of 5% for all agreements from not using a collection device, demonstrating that collecting less volume and using a designated saliva collection device was more effective. The use of a collection device improved the quality of saliva collected that was visually less viscous, less particulate, and uniformly clear-colored, leading to less accumulation of potential inhibitors.

It is also important to always consider handling precautions during the collection and processing of saliva via heat treatment alone, as the presence of contaminants may inhibit the RT-qPCR reaction. This includes avoiding powdered gloves during collection, transport, and handling, as they are common PCR inhibitors (Lomas et al., 1992; Broyles et al., 2002). Good laboratory practices recommend the use of nitrile gloves in a molecular laboratory (Viana and Wallis).

One of the highlights of our study is that we also monitored the stability of saliva specimens and found it remained stable at cold (2-8°C) and room temperature in the laboratory (∼25°C) for up to 5 days. Our finding is concordant with previous studies done by spiking SARS-CoV-2 into saliva specimens (Williams et al., 2021). SARS-CoV-2 may remain stable in saliva despite the presence of RNases in the medium due to its hard outer shell (Goh et al., 2020). This finding is promising for the utilization of saliva specimens in vast countries like Indonesia, where sending specimens from remote areas to diagnostic centers could take days. Nevertheless, we recommend using iceboxes during the transportation of saliva specimens as outside temperature during the day could be hotter than 30°C (WEATHER AND CLIMATE IN INDONESIA | Facts and Details). Once arrived at the laboratory, saliva specimens should also be processed as soon as possible. The sooner the diagnosis is made for the suspected patient; the sooner subsequent measures could be taken, hence limiting the transmission of the disease.

This method could also be used for serial screening of workplaces or schools as frequent NPOP swabbing causes discomfort. The 97% overall agreement of this method is superior compared to the antigen swab (Mak et al., 2020; Scohy et al., 2020). The developed method of treatment is versatile for several commercial SARS-CoV-2 RT-qPCR kits available in Indonesia. Although there was no particular association observed between the compatibility of saliva undergoing RNA-extraction-free treatment and kits’ characteristics, we demonstrated that this method could be adopted with several commercial kits. This allows the implementation of minimally treated saliva specimens across laboratories utilizing various commercial RT-qPCR kits without disrupting their existing workflows, with prior validation of their compatibility highly recommended.

The limitation of this study is that we did not retest the invalid samples for confirmation. Expert guidelines recommend repeating the test process when samples return an invalid result, either with the same workflow or, if available, using an alternate RT-qPCR kit to avoid reporting invalid (Perhimpunan Dokter Spesialis Patologi Klinik dan Kedokteran Laboratorium Indonesia, 2020). This process of retesting is highly recommended to be implemented with treated saliva as well. Hence a future study on this would elucidate the cause of discordance between specimen types.

In conclusion, we validated the use of human saliva as a viable alternative specimen to detect SARS-CoV-2 via direct RT-qPCR. Saliva can be collected in a tube without additives and remained stable at cold and room temperature for five days of storage. Upon arrival in the laboratory, saliva specimens can be treated against the heat incubation method alone, followed by immediate addition as a template for RT-qPCR reaction (**Figure 4**).

**Figure 4.**
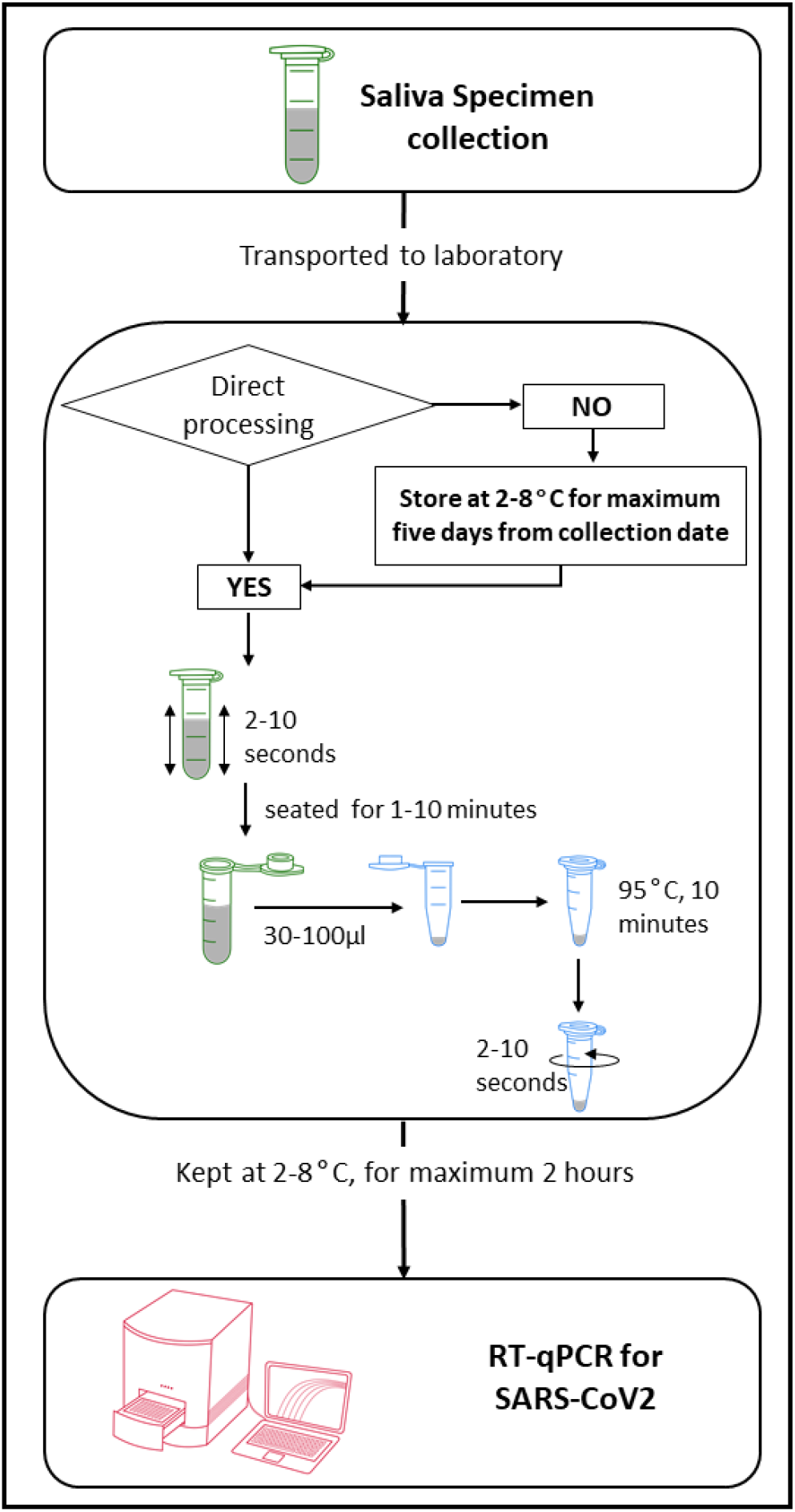
**Schematic workflow for collection and treatment of human saliva specimen for RNA-extraction free detection of SARS-CoV-2 via RT-qPCR.**

## Data Availability

Experimental results generated or analyzed during this study are included in this article and supplementary information. Experimental raw data are available upon request. Participants' information and data gathered through the informed consent and questionnaires are restricted to authors.

## 6 Author contributions

Contributed to study design: CM, SA, MMMK, SRV. Contributed to patient recruitment: SRV, SSS, MMMK, ET, FC, KIJ. Performed laboratory experiments: SRV, TAW, MMMK. Contributed to patient data management: SA, CM. Data analysis: CM, SRV, MMMK, SA. Manuscript writing: CM, SRV, SA, MMMK. Supervised and conceived the study: CM, SA, AI. All authors have reviewed the manuscript.

## 8 Acknowledgments

We would like to thank all the participating healthcare personnel and volunteers of our collaborating sites for contributing to specimen collection: FKIK Atma Jaya, Rumah Sakit Atma Jaya, Rumah Sakit EMC Tangerang, Intibios Lab Mangga Besar, Intibios Lab Pantai Indah Kapuk, and PT Nalagenetik Riset Indonesia. We would also like to acknowledge all patients and respondents who have voluntarily provided their specimen for this study, Sheila Jonnatan for her assistance in the early stage of the laboratory work, COVID-19 Laboratory (AJCUI) for providing the different RT-qPCR kits, and members of PT Nalagenetik Riset Indonesia in assistance for QuickSpit™ design and development. This work was supported by the COVID-19 Laboratory Center of Atma Jaya Catholic University of Indonesia (AJCUI).

## SUPPLEMENTARY INFORMATION

**Supplementary 1.** Optimization of RNA-Extraction-free treatment of saliva specimens.

| Treatment | Sample type | Extraction |
| --- | --- | --- |
| 1 | NPOP | Qiagen |
| 2 | Saliva | Qiagen |
| 3 | Saliva | 1.Transfer 100µl<br>2. Add 10ul Proteinase K<br>3. Incubate sample at 37°C for 30 mins<br>4. Heat inactivation at 95°C for 10 mins |
| 4 | Saliva | 1. Transfer 100µl<br>2. Add 10ul Proteinase K<br>3. Incubate sample at 37°C for 15 mins<br>4. Heat inactivation at 95°C for 10 mins |
| 5 | Saliva | 1. Transfer 100µl<br>2. Incubate sample at 37°C for 30 mins<br>3. Heat inactivation at 95°C for 10 mins |
| 6 | Saliva | 1. Transfer 100µl<br>2. Heat inactivation at 95°C for 10 mins |
| 7 | Saliva | 1. Transfer 100µl<br>2. Heat inactivation at 95°C for 10 mins<br>3. Centrifuge at max speed (13200 rpm) for 2 mins |
| 8 | Saliva | 1. Transfer 100µl<br>2. Heat inactivation at 95°C for 10 mins<br>3. Centrifuge at max speed (13200 rpm) for 5 mins |

**Supplementary 2.**
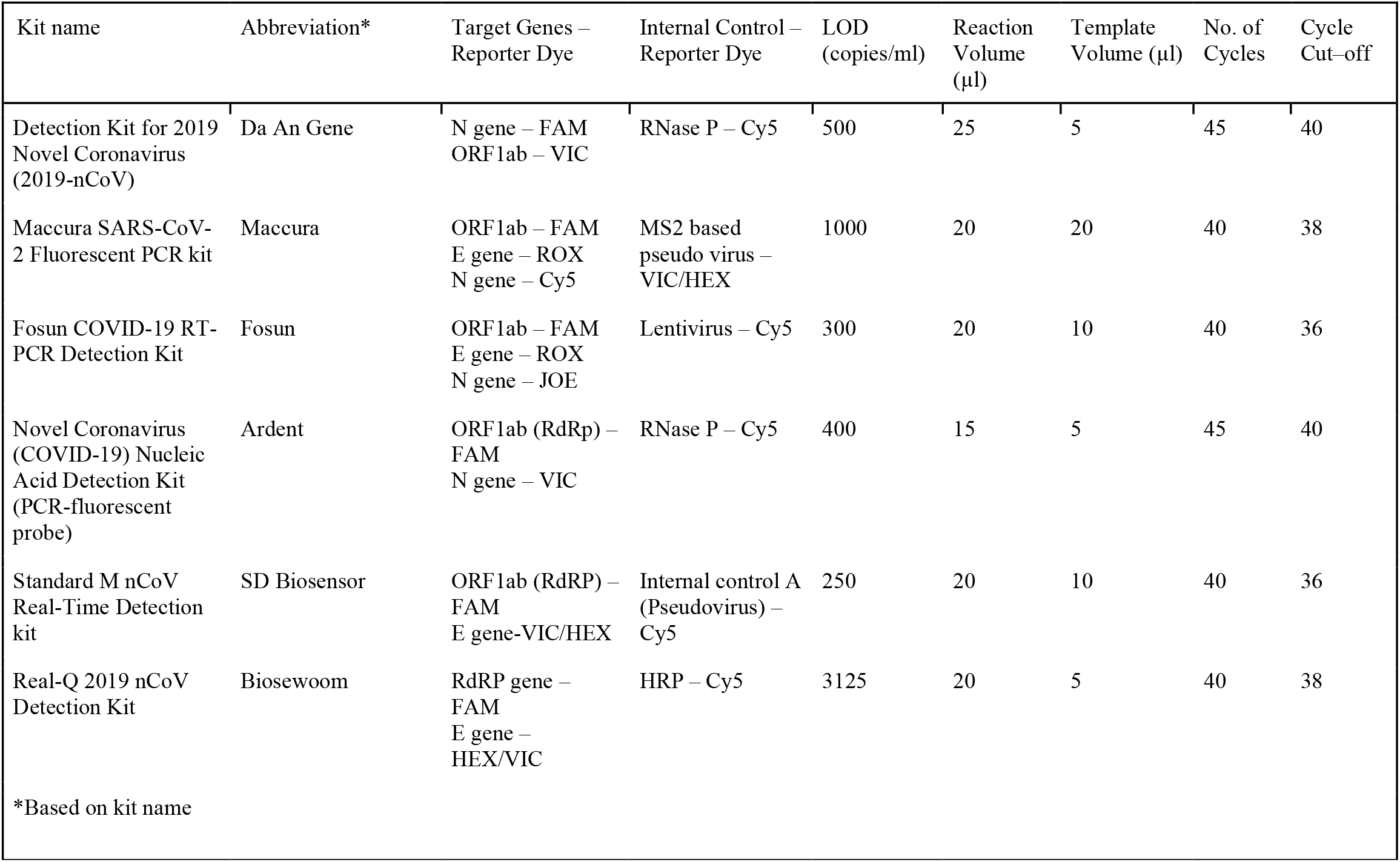
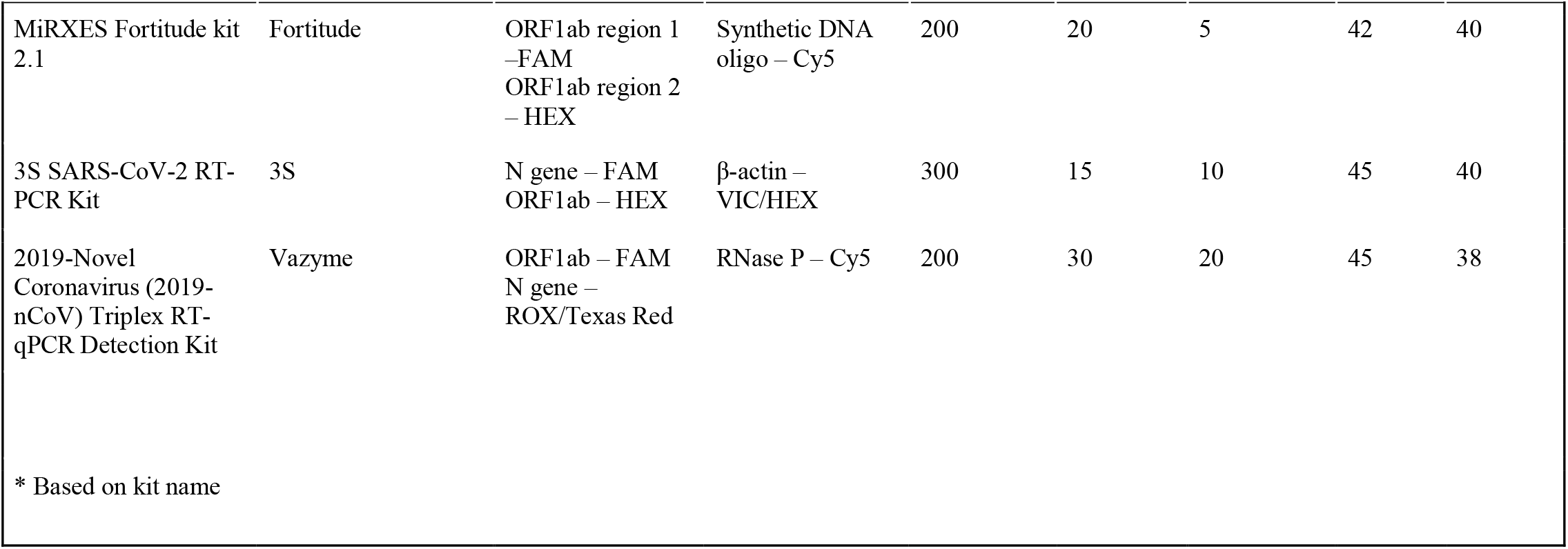
Specifications and characteristics of the commercial RT-qPCR kits, as derived from their respective manufacturer’s instructions.

**instructions**
| Kit name | Abbreviation* | Target Genes – Reporter Dye | Internal Control – Reporter Dye | LOD (copies/ml) | Reaction Volume (µl) | Template Volume (µl) | No. of Cycles | Cycle Cut-off |
| --- | --- | --- | --- | --- | --- | --- | --- | --- |
| Detection Kit for 2019 Novel Coronavirus (2019-nCoV) | Da An Gene | N gene – FAM<br>ORF1ab – VIC | RNase P – Cy5 | 500 | 25 | 5 | 45 | 40 |
| Maccura SARS-CoV-2 Fluorescent PCR kit | Maccura | ORF1ab – FAM<br>E gene – ROX<br>N gene – Cy5 | MS2 based pseudo virus – VIC/HEX | 1000 | 20 | 20 | 40 | 38 |
| Fosun COVID-19 RT-PCR Detection Kit | Fosun | ORF1ab – FAM<br>E gene – ROX<br>N gene – JOE | Lentivirus – Cy5 | 300 | 20 | 10 | 40 | 36 |
| Novel Coronavirus (COVID-19) Nucleic Acid Detection Kit (PCR-fluorescent probe) | Ardent | ORF1ab (RdRp) – FAM<br>N gene – VIC | RNase P – Cy5 | 400 | 15 | 5 | 45 | 40 |
| Standard M nCoV Real-Time Detection kit | SD Biosensor | ORF1ab (RdRP) – FAM<br>E gene-VIC/HEX | Internal control A (Pseudovirus) – Cy5 | 250 | 20 | 10 | 40 | 36 |
| Real-Q 2019 nCoV Detection Kit | Biosewoom | RdRP gene – FAM<br>E gene – HEX/VIC | HRP – Cy5 | 3125 | 20 | 5 | 40 | 38 |
| *Based on kit name |  |  |  |  |  |  |  |  |

| Kit name | Abbreviation* | Target Genes – Reporter Dye | Internal Control – Reporter Dye | LOD (copies/ml) | Reaction Volume (μl) | Template Volume (μl) | No. of Cycles | Cycle Cut-off |
| --- | --- | --- | --- | --- | --- | --- | --- | --- |
| MiRXES Fortitude kit 2.1 | Fortitude | ORF1ab region 1 –FAM<br>ORF1ab region 2 – HEX | Synthetic DNA oligo – Cy5 | 200 | 20 | 5 | 42 | 40 |
| 3S SARS-CoV-2 RT-PCR Kit | 3S | N gene – FAM<br>ORF1ab – HEX | β-actin – VIC/HEX | 300 | 15 | 10 | 45 | 40 |
| 2019-Novel Coronavirus (2019-nCoV) Triplex RT-qPCR Detection Kit | Vazyme | ORF1ab – FAM<br>N gene – ROX/Texas Red | RNase P – Cy5 | 200 | 30 | 20 | 45 | 38 |
| * Based on kit name |  |  |  |  |  |  |  |  |

**Supplementary 3.**
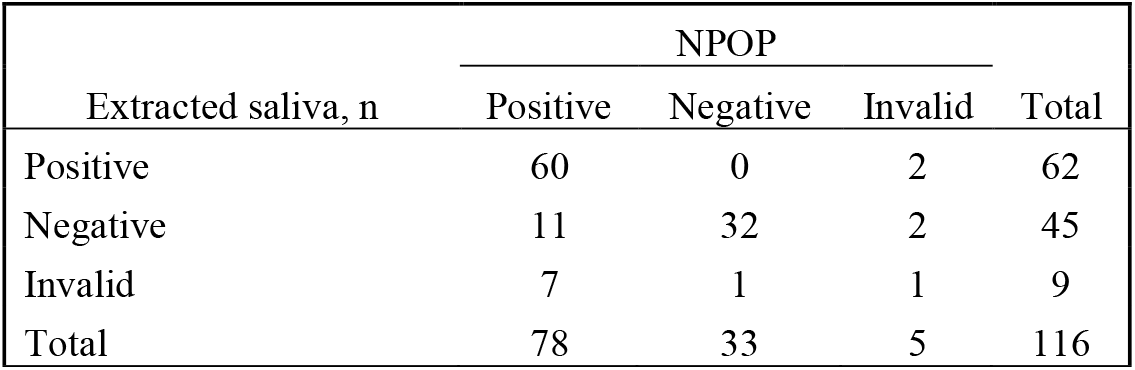
SARS-CoV-2 detection from RNA extracted from NPOP *versus* saliva specimens.

**Supplementary 4.**
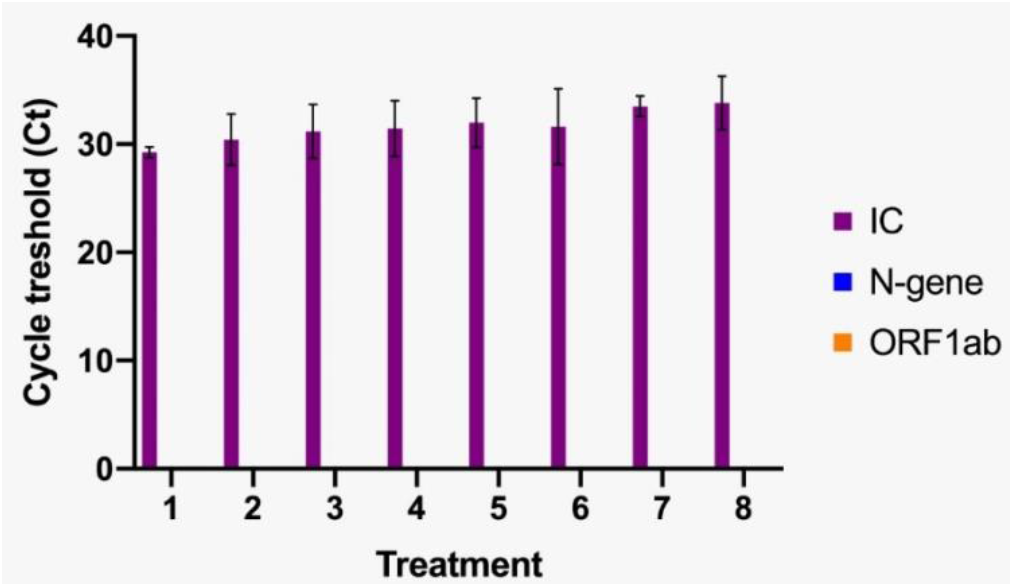
Comparison of eight different sample treatments prior to SAR-CoV-2 detection via PCR for negative saliva specimens. Bar graphs represent means ± SEM for three SARS-CoV-2 negative specimens tested for each sample treatment illustrated in **Figure 1A**.

**Supplementary 5.**
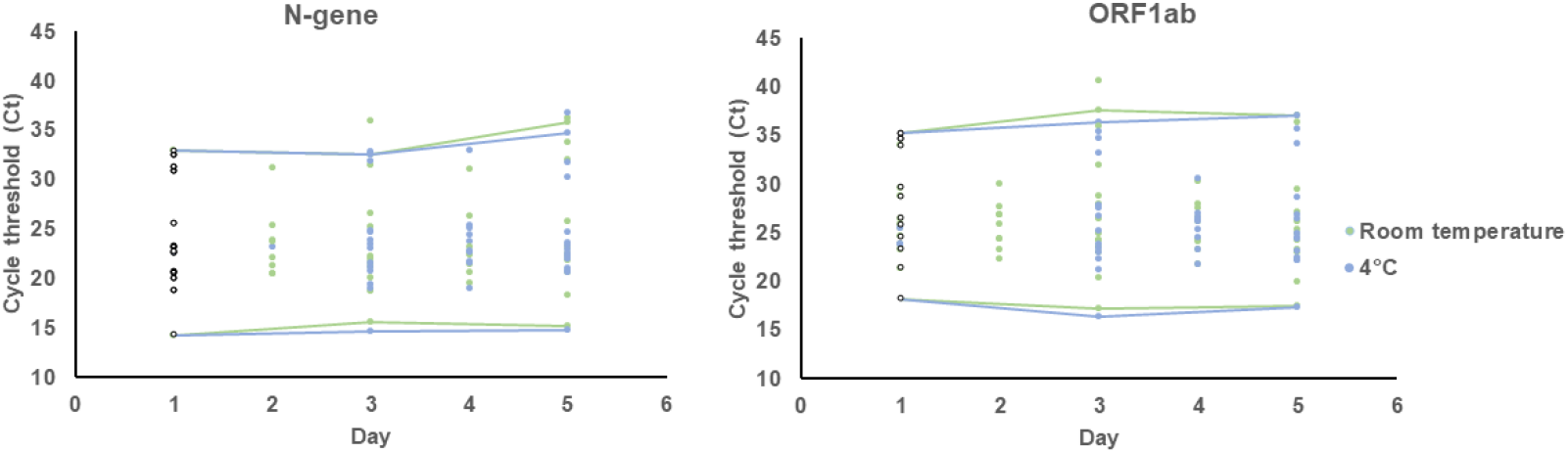
Detection of SARS-CoV-2 target genes N and ORF1ab plotted as the Ct value obtained for monitored sample. Data points collected in Day 1 are displayed as white circles with black outlines. Blue points refer to samples stored at cold while green at room temperature. Straight lines are drawn monitor changes for samples starting with the highest and lowest Ct value in Day 1.

**Supplementary 6.** Validation as RNA-extraction-free treatment of saliva.

| Treated saliva, n | NPOP |  |  | Total |
| --- | --- | --- | --- | --- |
|  | Positive | Negative | Invalid |  |
| Positive | 57 | 0 | 1 | 58 |
| Negative | 15 | 38 | 3 | 56 |
| Invalid | 9 | 1 | 1 | 11 |
| Total | 81 | 39 | 5 | 125 |

**Supplementary 7.** Performance of commercial kits using treated saliva as template for RT-qPCR.

| Commercial kit | Sample size, n | True positive, n | False positive, n | True negative, n | False negative, n | Invalid samples, n | Invalid rate, % | Overall agreement, % | Estimated sensitivity, % |
| --- | --- | --- | --- | --- | --- | --- | --- | --- | --- |
| Maccura | 11 | 11 | 0 | 0 | 0 | 0 | 0.00 | 100.00 | 100.00 |
| Fosun | 12 | 6 | 0 | 5 | 0 | 1 | 8.33 | 100.00 | 100.00 |
| Ardent | 11 | 10 | 0 | 0 | 1 | 0 | 0.00 | 90.91 | 90.91 |
| SD Biosensor | 13 | 8 | 0 | 3 | 1 | 1 | 7.69 | 91.67 | 88.89 |
| Biosewoom | 11 | 8 | 0 | 0 | 2 | 1 | 9.09 | 80.00 | 80.00 |
| Fortitude | 11 | 7 | 0 | 0 | 3 | 1 | 9.09 | 70.00 | 70.00 |
| 3S | 10 | 2 | 6 | 2 | 0 | 0 | 0.00 | 40.00 | 100.00 |
| Vazyme | 10 | 2 | 4 | 0 | 0 | 4 | 40.00 | 33.33 | 100.00 |

**Supplementary 8.** Implementation of QuickSpit™ as a device for collection of saliva specimen.

| Treated saliva collected with QuickSpit™, n | NPOP |  |  | Total |
| --- | --- | --- | --- | --- |
|  | Positive | Negative | Invalid |  |
| Positive | 45 | 0 | 0 | 45 |
| Negative | 8 | 240 | 2 | 250 |
| Invalid | 10 | 1 | 0 | 11 |
| Total | 63 | 241 | 2 | 306 |

**Supplementary 9.** Calculation of invalid samples in the different specimens and treatment conditions.

|  | Extracted NPOP | Extracted saliva | Treated saliva | Treated saliva collected with QuickSpit™ |
| --- | --- | --- | --- | --- |
| valid | 535 | 107 | 114 | 295 |
| invalid | 29 | 9 | 11 | 11 |
| total, n | 564 | 116 | 125 | 306 |
| % invalid | 5.14 | 7.76 | 8.80 | 3.59 |

## References

Alcoba-Florez, J., González-Montelongo, R., Íñigo-Campos, A., de Artola, D. G.-M., Gil-Campesino, H., Team, T. M. T. S., et al. (2020). Fast SARS-CoV-2 detection by RT-qPCR in preheated nasopharyngeal swab samples. Int. J. Infect. Dis. 97, 66–68. doi:10.1016/j.ijid.2020.05.099.

Azzi, L., Carcano, G., Gianfagna, F., Grossi, P., Gasperina, D. D., Genoni, A., et al. (2020). Saliva is a reliable tool to detect SARS-CoV-2. J. Infect. 81, e45–e50. doi:10.1016/j.jinf.2020.04.005.

Brian, D. A., and Baric, R. S. (2005). Coronavirus genome structure and replication. Curr. Top. Microbiol. Immunol. 287, 1–30. doi:10.1007/3-540-26765-4_1.

Broyles, J. M., O’Connell, K. P., and Korniewicz, D. M. (2002). PCR-based method for detecting viral penetration of medical exam gloves. J. Clin. Microbiol. 40, 2725–2728. doi:10.1128/jcm.40.8.2725-2728.2002.

COVID-19 developments in Indonesia.

Goh, G. K.-M., Dunker, A. K., Foster, J. A., and Uversky, V. N. (2020). Rigidity of the Outer Shell Predicted by a Protein Intrinsic Disorder Model Sheds Light on the COVID-19 (Wuhan-2019-nCoV) Infectivity. Biomolecules 10. doi:10.3390/biom10020331.

Griesemer, S. B., van Slyke, G., Ehrbar, D., Strle, K., Yildirim, T., Centurioni, D. A., et al. (2020). Evaluation of specimen types and saliva stabilization solutions for SARS-CoV-2 testing. medRxiv. doi:10.1101/2020.06.16.20133041.

Hung, K.-F., Sun, Y.-C., Chen, B.-H., Lo, J.-F., Cheng, C.-M., Chen, C.-Y., et al. (2020). New COVID-19 saliva-based test: How good is it compared with the current nasopharyngeal or throat swab test? J. Chin. Med. Assoc. 83, 891–894. doi:10.1097/JCMA.0000000000000396.

Iwasaki, S., Fujisawa, S., Nakakubo, S., Kamada, K., Yamashita, Y., Fukumoto, T., et al. (2020). Comparison of SARS-CoV-2 detection in nasopharyngeal swab and saliva. J. Infect. 81, e145– e147. doi:10.1016/j.jinf.2020.05.071.

Landry, M. L., Criscuolo, J., and Peaper, D. R. (2020). Challenges in use of saliva for detection of SARS CoV-2 RNA in symptomatic outpatients. J. Clin. Virol. 130. doi:10.1016/j.jcv.2020.104567.

Lomas, J., Sunzeri, F., and Busch, M. (1992). False-negative results by polymerase chain reaction due to contamination by glove powder. Transfusion 32, 83–85. doi:10.1046/j.1537-2995.1992.32192116439.x.

Mak, G. C., Cheng, P. K., Lau, S. S., Wong, K. K., Lau, C. S., Lam, E. T., et al. (2020). Evaluation of rapid antigen test for detection of SARS-CoV-2 virus. J. Clin. Virol. 129, 104500. doi:10.1016/j.jcv.2020.104500.

Ott, I. M., Strine, M. S., Watkins, A. E., Boot, M., Kalinich, C. C., Harden, C. A., et al. Simply saliva: stability of SARS-CoV-2 detection negates the need for expensive collection devices. doi:10.1101/2020.08.03.20165233.

Perhimpunan Dokter Spesialis Patologi Klinik dan Kedokteran Laboratorium Indonesia (2020). PANDUAN TATALAKSANA PEMERIKSAAN TES CEPAT MOLEKULER (TCM) dan POLYMERASE CHAIN REACTION (PCR) SARS-CoV-2. Website. Available at: https://www.pdspatklin.or.id/assets/files/pdspatklin_2020_04_30_19_20_35.pdf [Accessed May 25, 2021].

Scohy, A., Anantharajah, A., Bodéus, M., Kabamba-Mukadi, B., Verroken, A., and Rodriguez-Villalobos, H. (2020). Low performance of rapid antigen detection test as frontline testing for COVID-19 diagnosis. J. Clin. Virol. 129, 104455. doi:10.1016/j.jcv.2020.104455.

Smyrlaki, I., Ekman, M., Lentini, A., Rufino de Sousa, N., Papanicolaou, N., Vondracek, M., et al. (2020). Massive and rapid COVID-19 testing is feasible by extraction-free SARS-CoV-2 RT-PCR. Nat. Commun. 11, 4812. doi:10.1038/s41467-020-18611-5.

Timeline of WHO’s response to COVID-19.

Viana, R. V, and Wallis, C. L. 3 Good Clinical Laboratory Practice (GCLP) for Molecular Based Tests Used in Diagnostic Laboratories.

Vogels, C. B. F., Watkins, A. E., Harden, C. A., Brackney, D. E., Shafer, J., Wang, J., et al. (2020). SalivaDirect: A Simplified and Flexible Platform to Enhance SARS-CoV-2 Testing Capacity. Med. doi:10.1016/j.medj.2020.12.010.

Wang, W., Xu, Y., Gao, R., Lu, R., Han, K., Wu, G., et al. (2020). Detection of SARS-CoV-2 in Different Types of Clinical Specimens. JAMA 323, 1843–1844. doi:10.1001/jama.2020.3786.

Watkins, A. E., Fenichel, E. P., Weinberger, D. M., Vogels, C. B. F., Brackney, D. E., Casanovas-Massana, A., et al. (2020). Pooling saliva to increase SARS-CoV-2 testing capacity. medRxiv. doi:10.1101/2020.09.02.20183830.

WEATHER AND CLIMATE IN INDONESIA | Facts and Details.

Williams, E., Isles, N., Chong, B., Bond, K., Yoga, Y., Druce, J., et al. (2021). Detection of SARS-CoV-2 in saliva: implications for specimen transport and storage. J. Med. Microbiol. 70. doi:10.1099/jmm.0.001285.

World Health Organization (2020). Diagnostic testing for SARS-CoV-2. Available at: https://www.who.int/publications/i/item/diagnostic-testing-for-sars-cov-2.

Wyllie, A. L., Fournier, J., Casanovas-Massana, A., Campbell, M., Tokuyama, M., Vijayakumar, P., et al. (2020). Saliva or Nasopharyngeal Swab Specimens for Detection of SARS-CoV-2. N. Engl. J. Med. 383, 1283–1286. doi:10.1056/nejmc2016359.

Xu, R., Cui, B., Duan, X., Zhang, P., Zhou, X., and Yuan, Q. (2020). Saliva: potential diagnostic value and transmission of 2019-nCoV. Int. J. Oral Sci. 12, 11. doi:10.1038/s41368-020-0080-z.

Zhang, T., Cui, X., Zhao, X., Wang, J., Zheng, J., Zheng, G., et al. (2020). Detectable SARS-CoV-2 viral RNA in feces of three children during recovery period of COVID-19 pneumonia. J. Med. Virol. 92, 909–914. doi:10.1002/jmv.25795.

